# GDBIG: the first birth cohort genomic database and platform facilitating intergenerational genetic research

**DOI:** 10.1101/2024.12.27.24319711

**Authors:** Shujia Huang, Chengrui Wang, Mingxi Huang, Jinhua Lu, Jian-Rong He, Shanshan Lin, Siyang Liu, Huimin Xia, Xiu Qiu

## Abstract

High-quality genome databases derived from large-scale family-based birth cohorts are essential resources for investigating genetic determinants affecting early-life traits and the impact of early-life environments on the health of both parents and offspring. Here, we have established the genome database of Born in Guangzhou Cohort Study (BIGCS), name as GDBIG, which represents the first birth-cohort-based genomic database in China and is designed to facilitate generational genetic research, based on the Phase I results of BIGCS, that contains the low-coverage (∼6.63x) whole-genome sequencing (WGS) data and numerous pregnancy phenotypes of 4,053 Chinese participants. The participants were from 30 out of 34 administrative divisions of China, covering Han and 12 minority ethnic groups. Currently, GDBIG provides a comprehensive range of services, including allele frequency inquiries for 56.23 million variants across two generations, a genotype imputation server featuring a high-quality family-based reference panel, and a GWAS meta-analysis interface for various maternal and infant phenotypes. The GDBIG database addresses the dearth of Asian birth-cohort-based genomic resources and provides a valuable platform for conducting genetic analysis online or through application programming interfaces at http://gdbig.bigcs.com.cn/.

## Introduction

Genetic variation plays a significant role in determining individual susceptibility to almost all human diseases[1]. Consequently, elucidating the relationship between sequence variation and disease predisposition is a powerful strategy for uncovering fundamental processes related to disease pathogenesis, treatment, and prevention. Recent advancements in sequencing technologies[2,3], analytical methodologies[4–7], and large-scale international and national genome sequencing initiatives[8–12] have significantly facilitated the identification of causal genes and variants. However, most genome sequencing projects to date have focused on unrelated adult individuals[8–12]. These efforts have yielded valuable online resources, such as TOPMed[11,13], ChinaMAP[14] NyuWa[15], and STROMICS[12], which provide platforms for querying variants and performing genome imputation. While these platforms have substantially advanced genetic research, their emphasis on unrelated individuals has constrained our understanding of the genetic determinants of traits manifesting early in life and the influence of early-life exposures on short- and long-term health outcomes for both parents and offsprings. To bridge these gaps, genome sequencing studies based on birth cohorts that recruit families in trios or duos, with longitudinal follow-up, are essential[16,17]. In recent years, several international and regional birth cohort studies have been initiated to investigate the complex interplay between genetic factors and early-life environmental influences in shaping lifelong health trajectories. These projects include those employing array genotyping strategies to assay participants, such as the Early Growth Genetics Consortium (EGG)[18], the Avon Longitudinal Study of Parents and Children (ALSPAC)[19], the Hyperglycemia and Adverse Pregnancy Outcome (HAPO) study[20] involving multi-ethnic populations, the Olmsted County Birth Cohort (OCBC)[21], the Norwegian Mother and Child Cohort Study (MoBa)[22], as well as the Danish National Birth Cohort (DNBC)[23,24], and those utilizing whole genome sequencing strategies, such as the Born in Guangzhou Cohort Study (BIGCS)[25] and the China National Birth Cohort (CNBC)[26]. A comprehensive genomic database reporting and providing utilities for genetic information from birth cohort genome studies would be immensely valuable, which not only enables genetic and genomic information as provided by the genomic studies of unrelated individuals[8–12], but also facilitate intergenerational research that cannot be addressed by genomic studies of unrelated individuals, especially for underrepresented populations like Asians. Despite the release of various birth cohort studies, such a comprehensive genetic database has not been established yet.

In this study, we present the Genome Database of Born in Guangzhou Cohort Study (GDBIG), the first birth cohort-based genomic database and platform, developed using data from the Born in Guangzhou Cohort Study (BIGCS), the largest-scale birth cohort study in Asia. BIGCS was initiated in 2012 in Guangzhou City, China and so far has recruited over 50,000 duo and trio families and deeply phenotyped the family participant from less than 20 weeks of gestation until the age of 18 years [25]. GDBIG aims to provide user-friendly access to genomic, genetic, and phenotypic analytical data and genomic utilities based on genome sequencing and phenotypic data from BIGCS. The current GDBIG database includes the phase I resource from BIGCS, encompassing 4,053 healthy individuals in duos and trios (332 trios, 1,406 duos, and 245 unrelated individuals) sequenced by whole genome sequencing. The database currently holds information on 56,230,613 bi-allelic variants, comprising 51,052,456 single nucleotide polymorphisms (SNPs) and 5,178,157 insertions-deletions polymorphisms (Indels).

The primary functions of the current GDBIG are as follows: Firstly, it provides a query interface that allows users to search for overall and provincial-level allele frequency of genetic variation detected through whole genome sequencing of BIGCS participants, along with visualization and genomics API query tools. Secondly, it offers a genotype imputation server based on a high-quality reference panel, ensuring more accurate genotype imputation for individuals of similar ancestry due to the involvement of family members, which enables precise long-range phasing. Lastly, it provides a GWAS meta-analysis interface for numerous maternal and infant phenotypes cataloged based on the GWAS analysis efforts carried out in BIGCS. Users can access and analyze the data online through the website (http://gdbig.bigcs.com.cn/) or via the genomic API (https://github.com/BIGCS-Lab/GDBIGtools).

## Data collection and database construction

### Data collection and processing

The GDBIG was established using data from BIGCS, which involved 4,053 healthy participants, including 332 trios (comprising fathers, mothers and offspring), 1,406 duos (consisting of 14 father-offspring and 1,392 mother-offspring pairs), as well as 245 unrelated individuals (70 children, 150 adult females and 25 adult males). All participants were recruited by the BIGCS program at the Guangzhou Women and Children’s Medical Center (GWCMC). The parents were aged between 20 to 45 years old, with 83.7% being mothers and 16.3% fathers. Among the children, the male-to-female ratio was 53.6% to 46.4%. The cohort represented diverse ethnicities and linguistic groups, including 98% Han Chinese and 12 minority ethnic groups, with participants originating from 30 of China’s 34 administrative divisions. Cantonese speakers constituted the predominant language group followed by Mandarin, Hakka and Min speakers (Figure S1; Table S1). We performed low-coverage whole genome sequencing (WGS) on whole blood samples from adults and cord blood samples from infants, achieving an average sequencing depth of ∼6.63x. Data preprocessing included the removal of poor-quality reads, followed by alignment and variant calling by applied GATK best practice joint calling protocol in these participants. After variant quality score recalibration (VQSR), stringent quality control filters were then applied. We remove all of the multi-allelic variants and further removed SNPs that were located within the low-complexity regions of GRCh38, leading to a final set of 56,230,613 biallelic variants, comprising 51,052,456 SNPs and 5,178,157 Indels. Among these variants, 22,276,474 (39.62%) were singleton (Allele count (AC) = 1) variants, 11,919,262 (21.20%) were doubletons (AC=2), 9,648,220 (17.16%) were very rare variants (AC>2 and AF ≤ 0.1%), 5,279,786 (9.39%) were rare variants (0.1%< AF ≤1%), 1,907,131 (3.39%) were low-frequency variants (1%< AF ≤5%), and 5,199,740 (9.25%) were common variants (AF > 5%). Approximately 32.56% of the variants (18,308,033 Ts/Tv=1.46) were not reported in NCBI dbSNP (Build 154), and 93.4% of were classified as singletons or doubletons (AC≤2). The variants were further annotated using Variant Effect Predictor (VEP) with default parameters[27]. Detailed information about the variant filtration processes can be found in our associated published paper[28].

### Database architecture and web server implementation

Based on the genomics data obtained from the BIGCS, we employed the following IT strategies to construct GDBIG. Data security was ensured through the implementation of a deployment scheme consisting of two servers, segregating the frontend application server from the backend server. This configuration restricted the transfer of genomics data solely through a controlled permission port within the local area network (LAN) between these servers (**Figure 1**).

**Figure 1.**
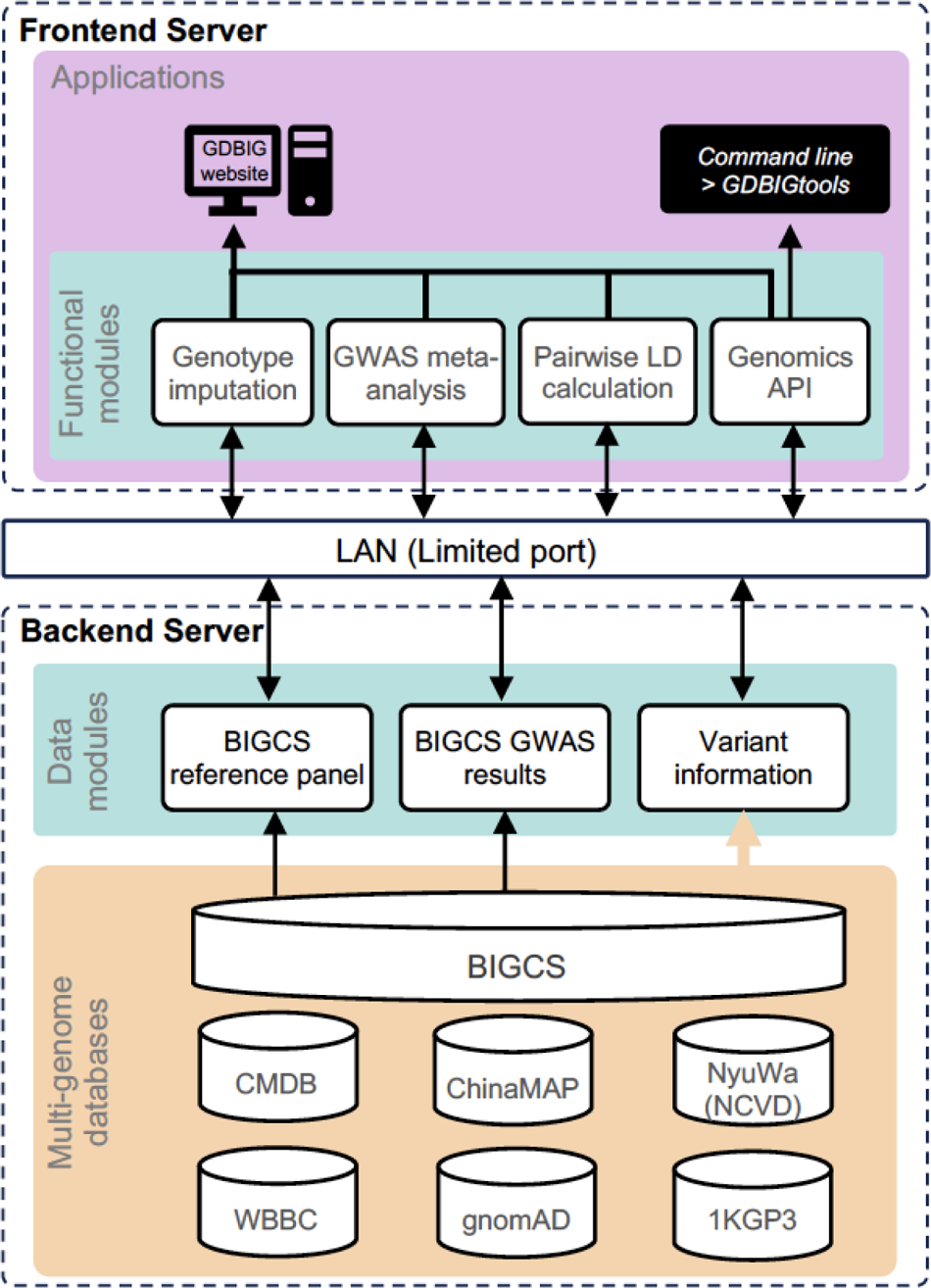
Architecture of the GDBIG website and database. The GDBIG website is divided into two parts: Frontend server and Backend server. The genomics data is stored exclusively in the Backend server and cannot be accessed directly from the Frontend. Data transfer is limited to a controlled permission port within the local area network (LAN). GDBIG consists of four functional modules, providing users with online access and a genomics API for variant searching, genotype imputation, GWAS meta-analysis, and Pairwise LD calculation through the GDBIG website interface on the Frontend server. In the Backend, the server stores variant information from BIGCS, BIGCS reference panel and BIGCS GWAS results. Additionally, six other human genome variation databases (CMDB, ChinaMAP, NyuWa, WBBC, gnomAD and 1KGP3) have been integrated into the Backend, facilitating allele frequency comparison with BIGCS dataset.

For the backend server, MongoDB (https://www.mongodb.com/) was utilized as the genome database engine. MongoDB is a widely used open-source document-oriented NoSQL database that is specifically designed for the storage of large-scale datasets. The application of MongoDB makes it convenient and secure to set up authentication for data access, updates or replenishment. In addition, GDBIG integrated variants data from various existing databases, including CMDB[29], ChinaMAP[30], NyuWa (NCVD)[15], WBBC[31], gnomAD[32] and 1KGP3[8]. This integration served as background databases, enabling cross-population comparisons of variation spectra, and supporting investigators in their research endeavors.

The frontend interface of GDBIG employed Gin as the web framework (https://gin-gonic.com/) and utilized MySQL for the management of user information and permissions (https://www.mysql.com/). Gin is a popular, free and open-source high-performance HTTP web framework written in Golang (Go, https://go.dev/). On the other hand, MySQL is the most widely used free relational database management system globally, well-suited for storing and managing user registration information. To visualize the data within GDBIG, the website utilized ECharts (https://echarts.apache.org/), a JavaScript library, and cross-platform open-source toolkit for data visualization. It is important to highlight that, due to the legal protection of the human genetic data stored in the database, researchers are required to provide basic registration and login information before accessing the retrieval and analysis modules.

## Database content and usage

### Searching and browsing variants on the website

The GDBIG browser provides access to variant information sourced from the GDBIG database. This information includes various details like chromosomal position, reference allele, mutated alleles, biological consequence, overall mutated allele frequencies, and the regional and provincial allele frequencies of the Chinese population in seven geographical areas (North, Northeast, Northwest, East, Central, Southwest, and South) for each variant. Users can access this information through the Search page on the GDBIG website (http://gdbig.bigcs.com.cn/), by queries with gene symbol, transcripts ID, RSID of variant, genomic region, or chromosomal positions. The website ensures rapid retrieval of variation information, which is further presented through clear visualization.

On the overview page, a dynamic display shows the summary information of BIGCS variants, allowing users to select specific chromosomes or zoom in/out on particular regions for quick access to variation details (**Figure 2A**). When searching for a genomic region or gene ID, the search result includes summary information, coverage read depth for each position, distribution of transcript location (if available), and annotated information on detected variants (**Figure 2B**). Moreover, the webpage provides selection buttons that enable users to filter variants based on functional types (missense, synonymous, loss of function (LoF)) or variant types (SNP, Indel) (**Figure 2C**, **D**).

**Figure 2.**
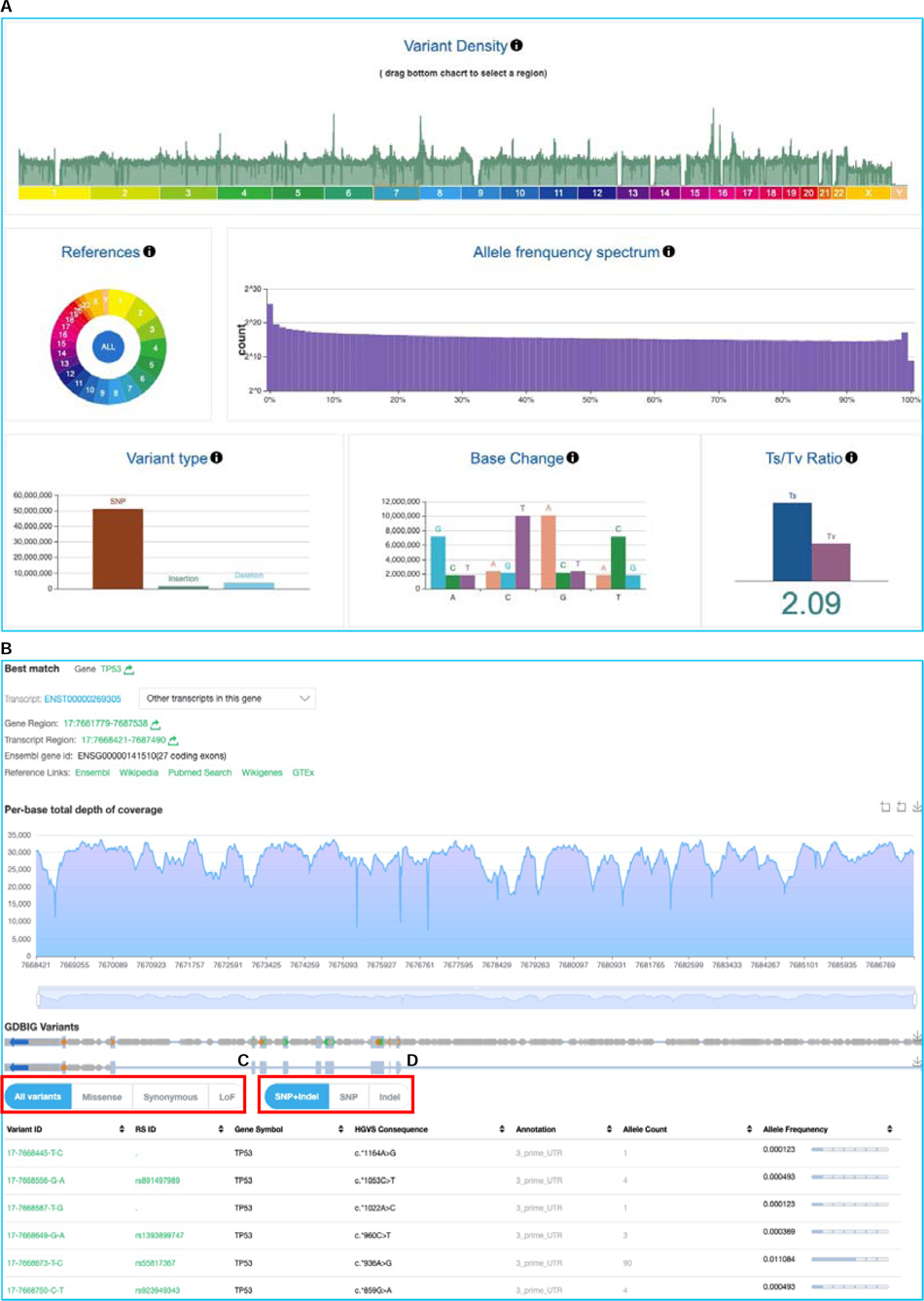
Distribution of variants and example of gene searching result in GDBIG. (A) Overview page presenting summary statistics of genomic variants in GDBIG. (B) Screenshot of gene search for *TP53*. The resulting variants within the *TP53* are listed below. Users can click on buttons to select specific functional annotations (C) or to choose different types of variants (SNP or Indel) (D).

Furthermore, by clicking on any of the search variants, users can access more detailed information, such as variant descriptions, allele frequency distributions across the seven geographical areas of China, and integrated variant information from worldwide populations available in existing databases like CMDB[33], ChinaMAP[30], NyuWa (NCVD)[15], WBBC[31], gnomAD[32], and 1KGP3[8]. This feature empowers users to investigate the frequency of the same variant from both a global and regional perspective (**Figure 3**), thereby proving invaluable for studying rare or localized diseases.

**Figure 3.**
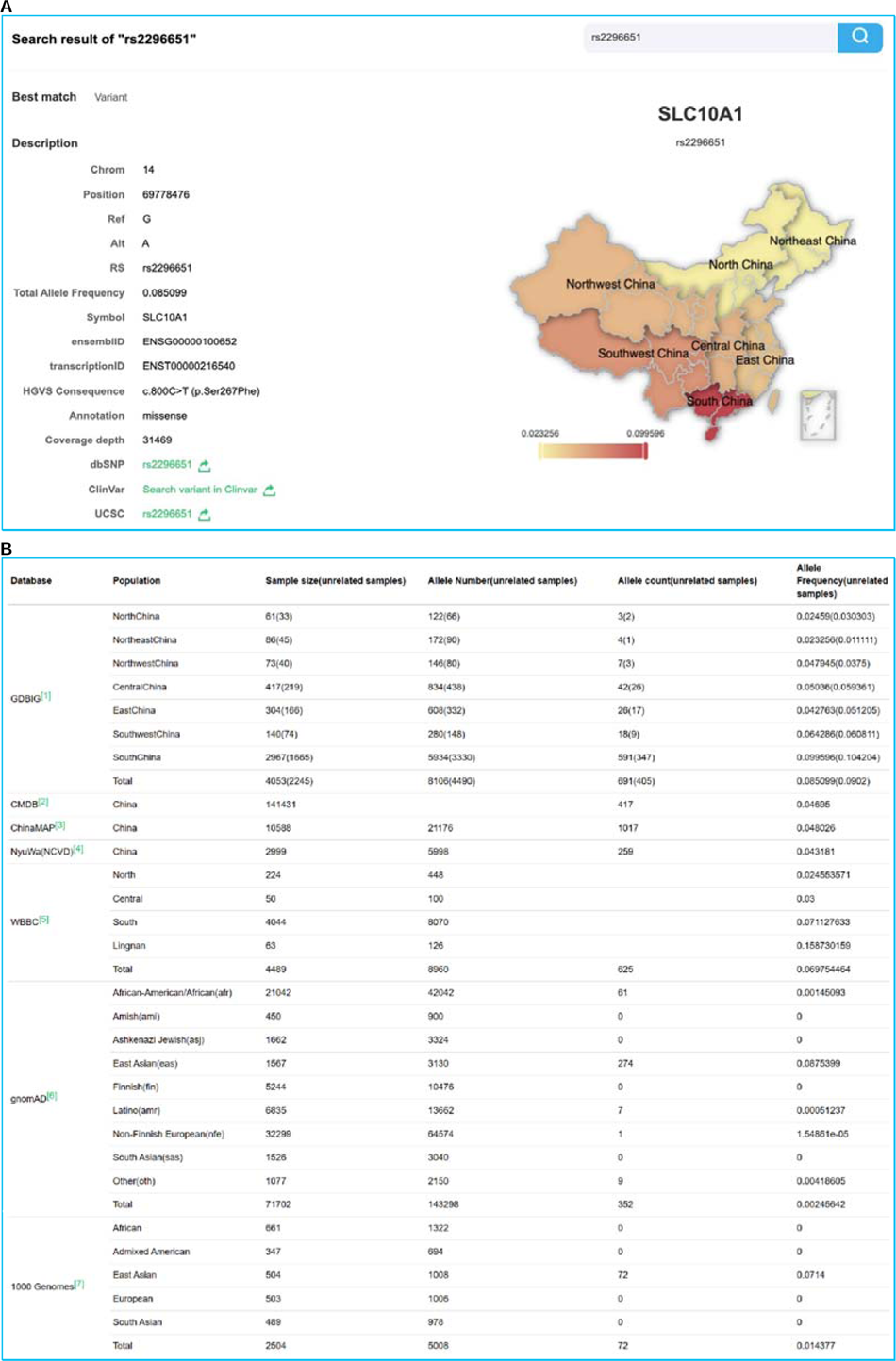
Example of variant searching result in GDBIG. (A) Search for the rs2296651 variant as an example. The search results display relevant information about the variant, including chromosomal position, allele information, allele frequency, gene ID, and the allele frequency in seven geographical areas of China (North, Northeast, Northwest, East, Central, Southwest and South). A geography map of China is provided on the righthand side, where the color intensity represents the allele frequency, with darker color indicating higher frequency. (B) Details allele frequency information for the variant in GDBIG compared to other genomic databases integrated into GDBIG.

### Genomics API and tools

In addition to the web search page and visualization tools, we recognize the importance of batch query functionality is essential for specific research objectives. To address this need, we have developed a genomic application programming interface (API) for GDBIG. This RESTful API (https://restfulapi.net/) allows researchers to remotely access variant data through program scripts or the command line.

GDBIGtools (https://github.com/BIGCS-Lab/GDBIGtools) has been developed based on the above GDBIG genomics API. It is a Python toolkit designed to facilitate variant queries and annotate local variation files in VCF format using information from GDBIG, such as overall allele frequency, allele frequency across different geographical areas, and filtration status, among other data (**Figure 1**). GDBIGtools is available on the Python Package Index (PyPI, https://pypi.org/) and can be easily installed using the Python PIP approach, by executing the command “pip install GDBIGtools” in the command line. Detailed instructions on the usage of GDBIGtools are provided on both the API ACCESS page (http://gdbig.bigcs.com.cn/api.html) and the corresponding Github repository. During practical application, GDBIGtools is capable of querying approximately 1,800 variants per minute, with performance influenced by the speed of the internet connection. The output of the query results is also provided in VCF format, ensuring compatibility and seamless integration with existing workflows.

### Genotype imputation server

Genotype imputation is a powerful method for accurately inferring individual genotypes at genomic positions not presented in arrays[34]. The application of genotype imputation with a population-specific reference panel can significantly enhance the power of GWAS studies[5,35–38]. In the context of BIGCS, the recruitment of family members in the study enables accurate long-range phasing and substantially improves the accuracy of genotype imputation[39]. The current reference panel was constructed using BIGCS Phase I participants’ data[28], leveraging unique family-relatedness information within the BIGCS cohort. To create the reference panel, we extracted all 2,245 unrelated individuals from the whole phased variant dataset mentioned above, which comprises 43,055,086 high-quality SNPs and 4,184,387 Indels, covering all 22 autosomes and the X chromosome. Notably, the reference panel includes 13.66 million novel variants (not present in dbSNP Build 154) and 30.88 million new variants previously absent in the 1KGP3 reference panel[8], which has been the primary reference panel for the Chinese population, containing 301 unrelated individuals of Chinese ancestry. To assess the utility of the BIGCS reference panel in imputing genotypes for individuals of Chinese ancestry, we employed an independent high-coverage WGS dataset comprising 50 Chinese individuals (40x coverage, 11,174,603 biallelic variants). We mimicked a standard imputation process using the 930,000 SNP sites from the Affymetrix Genome-Wide Human SNP Array 6.0 and estimated the imputation accuracy (*R^2^*) by comparing the imputed genotype dosage with true genotypes derived from high-coverage sequencing at sites absent on the array. Across all allele frequency ranges, the BIGCS panel consistently exhibited superior *R^2^* and true variant coverage when compared with commonly used reference panels for individuals of Chinese ancestry, including the 1KGP3 reference panel (n = 2,504), the haplotype reference consortium panel (HRC) (n = 32,470), the GenomeAsia100K Project reference panel (GAsP) (n = 1,654) and the multi-ethnic TOPMed reference panel (TOPMed) (n = 97,256). Specifically, BIGCS reference panel achieved an average *R*^2^ of 0.715 for low frequency variants (1% ≤ MAF < 5%) and 0.936 for common variants (MAF ≥ 5%)[28], outperforming four commonly used reference panels: 1KGP3[8] (low frequency: 0.687, common: 0.922), the haplotype reference consortium panel (HRC)[40] (low frequency: 0.660, common: 0.892), the GenomeAsia100K Project reference panel (GAsP)[41] (low frequency: 0.672, common: 0.873), and the multi-ethnic TOPMed reference panel (TOPMed)[11] (low frequency: 0.699, common: 0.920). For rare variants with MAF ≤ 1%, the BIGCS reference panel also exhibited the highest average *R*^2^ (*R*^2^ = 0.466) compared to the 1KGP3(*R*^2^=0.291), HRC(*R*^2^=0.206), GAsP(*R*^2^=0.240), and TOPMed(*R*^2^=0.422) panels. The higher imputation accuracy of the BIGCS reference panel unravels the value of addressing the underrepresentation of Chinese genetic diversity worldwide and highlights the benefits of a family design in constructing population haplotype references. More details about the BIGCS reference panel and its performance compared to these existing reference panels can be referred to our associated published paper[28]. For quality control standards in genotype imputation, we recommend that users sequence a subset of their samples at high depth and empirically evaluate the imputation accuracy by comparing the true genotypes with the imputed genotype dosages.

Within GDBIG, we have developed a dedicated web page and interface to facilitate genotype imputation based on the BIGCS reference panel. Users can access this tool under the “RESEARCH TOOLS” menu on the GDBIG website(http://gdbig.bigcs.com.cn/imputation_server/jobs.html). BEAGLE5[42] (https://faculty.washington.edu/browning/beagle/b5_1.html) with default parameters is employed for phasing process for the input VCF files and Minimac4 (https://genome.sph.umich.edu/wiki/Minimac4) is employed for imputation step with default parameters in “Imputation server”. The process is user-friendly and involves four steps (**Figure 4A**): (1) uploading a variants file in VCF format with coordinates conforming to GRCh38/hg38; (2) selecting the BIGCS reference panel and imputation tools; (3) submitting the job and awaiting its completion; (4) downloading the final results. Users will receive email notifications with detailed job status updates once the task is finished. On average, genotype imputation for an input VCF file containing 50 samples and 720k SNPs takes approximately one hour to complete (Table S2).

**Figure 4.**
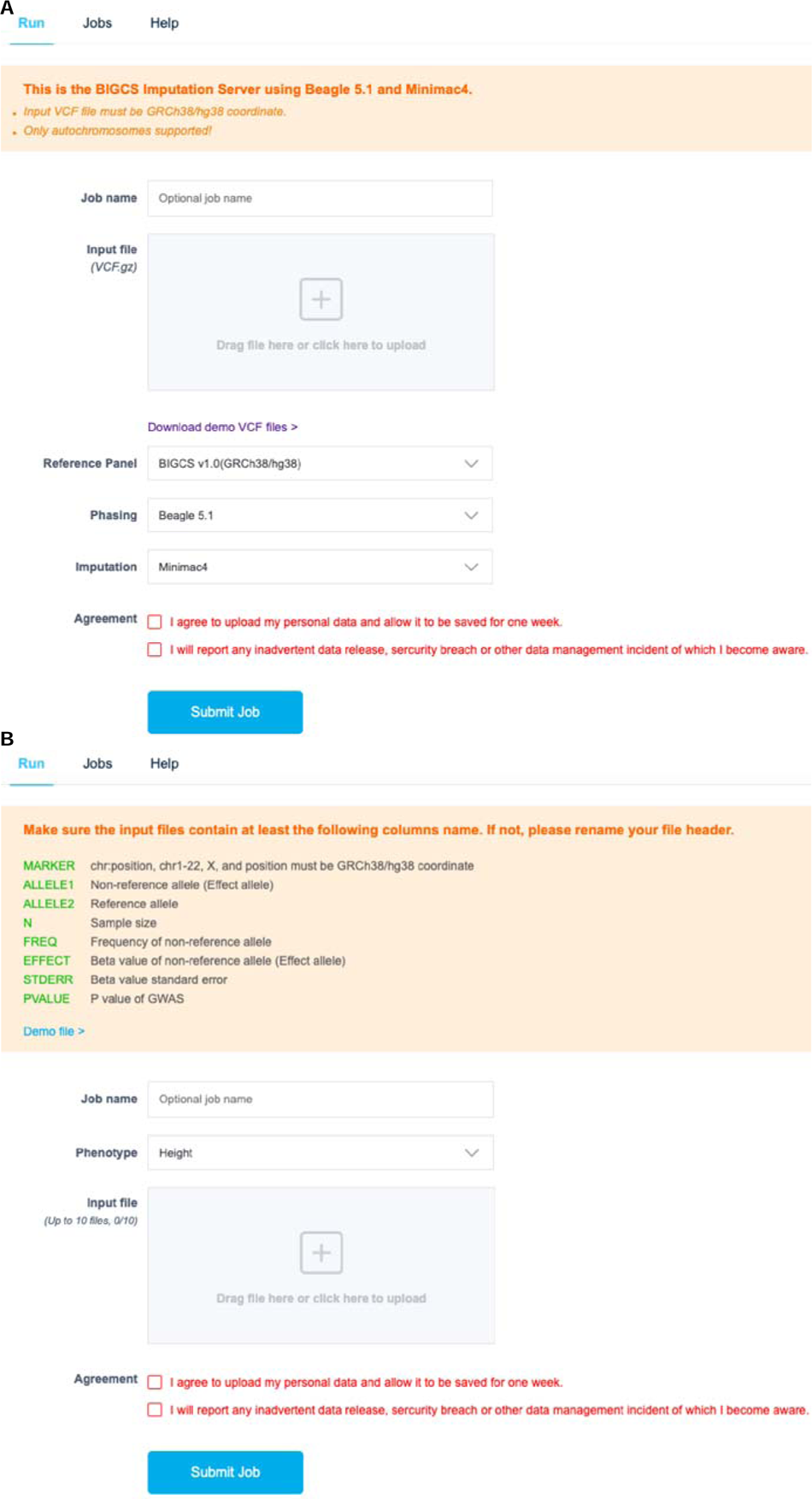
Web interface for genotype imputation (A) and GWAS meta-analysis (B).

### GWAS meta-analysis interface

GWAS meta-analysis has proven to be a valuable approach for enhancing the statistical power of association studies, particularly in cases with limited sample sizes. This method involves consolidating GWAS summary statistics from multiple studies for a large number of genetic variants[43–46]. To facilitate GWAS meta-analysis for general phenotypes as well as specific pregnancy and fetal growth phenotypes, GDBIG offers a dedicated portal and workflow that utilizes the METAL software[47].

In the initial phase of our BIGCS study, we conducted GWAS analyses on anthropometric, gestational, and fetal growth phenotypes. Currently, GDBIG provides GWAS summary statistics for 14 pregnancy-related phenotypes, encompassing adult anthropometric traits (height, weight, BMI), lipid metabolism traits during pregnancy (TBA, TC, HDL, LDL, TG), OGTT levels measured during weeks 24-28 of pregnancy (0H, 1H, 2H), gestational weight gain (GWG) rate (kg/week), and neonatal birth weight and birth length (**Table 1**). Genetic association signals for shared phenotypes in Chinese pregnancies sequenced through noninvasive prenatal testing (NIPT)[48] demonstrated a 100% replication rate[28]. The workflow for GWAS meta-analysis is designed to be as straightforward as the genotype imputation process, consisting of four steps (**Figure 4B**): (1) selecting the trait of interest; (2) uploading one or more GWAS summary statistics files corresponding to the selected trait; (3) submitting the jobs and awaiting completion; (4) downloading the meta-analysis results. Detailed instructions are available on the GWAS meta-analysis page (http://gdbig.bigcs.com.cn/gwas_meta_analysis/status.html). For quality control in meta-analyses, we recommend that users first evaluate genomic control measures, such as lambda values and quantile-quantile (Q-Q) plots[49], along with LD score regression metrics (intercept and ratio)[50] to assess genome-wide statistical inflation. Additionally, users are encouraged to perform replication analyses to validate the findings of the meta-GWAS.

**Table 1.** The adult and fetal growth phenotypes included in the GWAS meta-analysis module in GDBIG.

| **Traits** | **Sample size** | **Variants number** |
| --- | --- | --- |
| **Anthropometric** |  |  |
| Height (prepregnancy) | 2195 | 7,134,855 |
| Weight (prepregnancy) | 2165 | 7,131,603 |
| BMI (prepregnancy) | 2171 | 7,132,009 |
| Gestational weight gain (GWG) rate | 1566 | 7,110,421 |
| **Biochemical (Collection in 16-20 weeks of pregnancy)** |  |  |
| Total Bile Acid (TBA) of pregnancy | 1695 | 7,115,228 |
| Total cholesterol (TC) | 1535 | 7,112,498 |
| High-density lipoprotein (HDL) | 1532 | 7,111,976 |
| Low-density lipoprotein (LDL) | 1532 | 7,111,938 |
| Triglycerides (TG) | 1531 | 7,111,181 |
| **OGTT (Collection in 24-28 weeks of pregnancy)** |  |  |
| Glucose OGTT 0H of pregnancy | 1684 | 7,112,787 |
| Glucose OGTT 1H of pregnancy | 1687 | 7,112,953 |
| Glucose OGTT 2H of pregnancy | 1685 | 7,112,724 |
| **Fetal growth** |  |  |
| Birth Length | 1724 | 7107,435 |
| Birth Weight | 1746 | 7109,094 |

### Pairwise LD calculation based on BIGCS variant dataset

Additionally, we have included a module named "Pairwise LD calculation", which facilitates real-time linkage disequilibrium (LD) computations using the extensive BIGCS variants dataset. Access to this module can be obtained through the "RESEARCH TOOLS" menu on the GDBIG website (http://gdbig.bigcs.com.cn/ld/cal.html). Notably, unlike existing datasets, GDBIG offers LD calculations that include Indels, thereby addressing a previously unmet need in the field (**Figure 5**). To the best of our knowledge, this represents the first online tool specifically tailored to enable researchers to conduct LD calculations solely based on the Chinese population genetic data resource. Moreover, it successfully bridges the gap in LD calculation for Indels variants.

**Figure 5.**
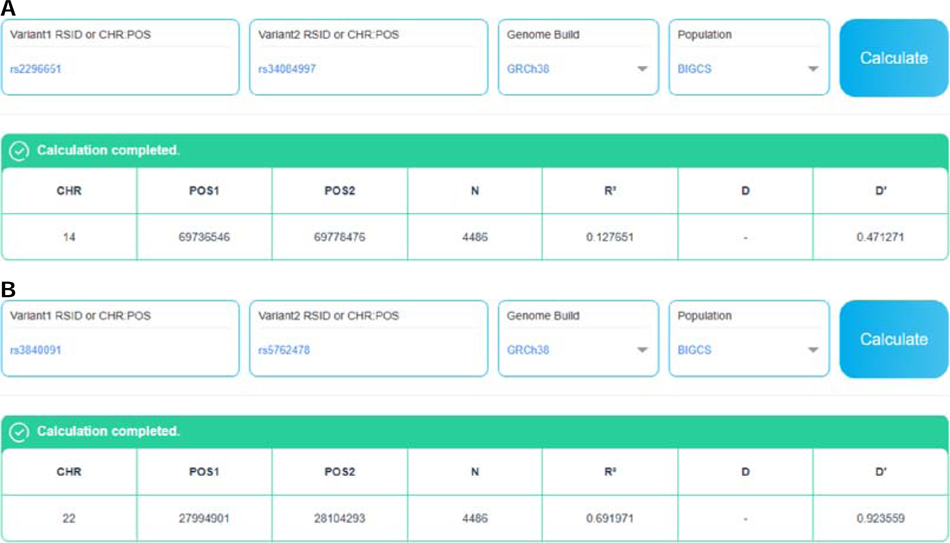
Examples of pairwise LD calculation. (A) Pairwise LD calculation between two SNPs: rs2296651(chr14-69778476-G-A) and rs34084997(chr14-69736546-C-T). (B) Pairwise LD calculation between an Indel (rs3840091, chr22-27994901-CCAGA-C) and a SNP (rs5762478, chr22-28104293-T-C).

## Discussion and perspectives

The release of GDBIG marks a noteworthy milestone as the first family-based genetic database and analysis platform associated with the largest birth cohort in China. It not only addresses the scarcity of large-scale maternal-infant whole-genome sequencing (WGS) variation datasets worldwide, especially for the underrepresented Asian population, but also provides a valuable platform for online analysis of population genetic data or utilization through the genomics API. The current release of GDBIG is founded on the WGS data of 4,053 individuals from Phase I BIGCS project. GDBIG encompasses a wide range of functionality, including the provision of functional annotations and global/regional allele frequency information for 56.23 million genomic variants. Additionally, it offers a high-quality reference panel with genotype imputation accuracy that outperforms other used Chinese reference panels. Furthermore, GDBIG provides a GWAS meta-analysis interface, presenting genotype-phenotype association results for fourteen pregnancy and fetal growth traits, among others. Moreover, GDBIG provides the online tool specifically tailored to enable researchers to conduct LD calculations solely based on the Chinese population genetic data resource, which based on the extensive BIGCS variants dataset. Importantly, GDBIG offers researchers a novel avenue for sharing and analyzing data, eliminating the need to disclose raw genetic data, thus ensuring data privacy. Overall, GDBIG constitutes a significant contribution to the field, providing researchers with an unprecedented resource for exploring and leveraging genomic data within the context of a birth cohort. The availability of such a comprehensive platform opens new avenues for understanding the genetic basis of various traits and diseases, thereby advancing genetic and genomic studies on a global scale.

Looking ahead, we aim to further develop and enhance the GDBIG by addressing its current limitations through the following endeavors: First, the current sample size for allele frequency inference, genotype imputation, and genome-wide association studies is limited to thousands of participants. As genomic data from the BIGCS cohort continues to expand, we anticipate augmenting the database by incorporating additional datasets and updating the variant catalog and BIGCS reference panel. Moreover, we intend to integrate more human genome variation databases from diverse global populations, thereby enhancing its comprehensiveness and representation. Second, the range of phenotypes covered by the database requires expansion. We will provide GWAS summary statistics for a wider spectrum of maternal pregnancy traits, fetal growth phenotypes, and birth defects, facilitating more extensive meta-analyses. With the anticipated increase in sample size from the first endeavor, the statistical power of these analyses will also be strengthened. Third, the database currently focuses on SNPs and Indels. Recognizing the critical role of structural variants (SVs) in the etiology of various genetic diseases and traits[51–53], we plan to incorporate SV data into the database. This effort will be supported by high-depth sequencing data from the Phase II BIGCS study, further enriching the database’s content and utility. Fourth, beyond the three primary analyses in GDBIG, we plan to deploy additional bioinformatics tools, such as Eagle[54] for haplotype phasing and genotype imputation, and MR-MEGA[55] for multi-ancestral meta-GWAS analysis. Moreover, we are developing interactive visualization tools for genomic data, which will be made available as online applications on the GDBIG platform. These tools aim to enhance the platform’s analytical capabilities and provide an improved user experience. Finally, we remain committed to improving the functionality and usability of GDBIGtools. Dedicated user support will be provided to assist researchers in utilizing the data and tools available within GDBIG effectively. Through these ongoing efforts, we strive to continuously advance and optimize GDBIG, ensuring it remains a robust, cutting-edge resource for genetic and intergenerational research.

## Ethical statement

The study was approved by the Ethics Committee of Guangzhou Women and Children’s Medical Center (no. 2012[015] and 2017102302). The participants consented to the publication of the research results.

## Data availability

The raw sequencing data in this work has been deposited at the National Genomics Data Center (NGDC; https://ngdc.cncb.ac.cn), China National Center for Bioinformation/ Beijing Institute of Genomics, Chinese Academy of Sciences, with accession number: HRA002496. Data can be accessed via applications, following the GSA guide (https://ngdc.cncb.ac.cn/gsa-human/document). All of the GWAS summary statistics data in this GDBIG platform has been deposited in the Genome Variation Map (GVM) in NGDC, under accession number GVP000003 (https://ngdc.cncb.ac.cn/gvm/getProjectDetail?project=GVP000003). The user can contact the corresponding author to apply for permission to access this data. In addition, the website of Genome Database of Born in Guangzhou Cohort Study (GDBIG) is freely available at http://gdbig.bigcs.com.cn/. Researchers can be accessed through an interactive API, which can be found at http://gdbig.bigcs.com.cn/api.html. Use tutorials are provided on the same webpage to assist users in navigating the platform effectively. The code for extracting information from GDBIG is available on the Github repository(https://github.com/BIGCS-Lab/GDBIGtools<u>) and</u> BioCode project at NGDC (https://ngdc.cncb.ac.cn/biocode/tool/7586). Variant dataset and the GWAS summary statistics for the 14 traits can also be accessed through the GDBIG server (http://gdbig.bigcs.com.cn/), with approval from the Human Genetic Resources Administration of China (HGRAC) (ID: 2022BAT1051). Researchers who are interesting in collaborating with the BIGCS group are encouraged to contact Xiu Qiu and.

## CRediT authorship contribution statement

**Shujia Huang:** Conceptualization, Investigation, Methodology, Project administration, Formal analysis, Software, Visualization, Validation, Writing - original draft, Writing - review & editing, Funding acquisition. **Chengrui Wang:** Investigation, Data curation, Formal analysis, Software, Visualization, Writing. **Mingxi Huang:** Investigation, Data curation, Formal analysis, Visualization, Validation. **Jinhua Lu:** Platform, Validation. **Jian-Rong He:** Validation, Writing - review & editing. **Shanshan Lin:** Validation, Writing - review & editing. **Siyang Liu:** Validation, Writing - review & editing. **Huimin Xia:** Funding acquisition, Resources. **Xiu Qiu:** Conceptualization, Supervision, Writing - review & editing, Funding acquisition, Resources, Validation, Project administration. All authors have read and approved the final manuscript.

## Data Availability

The raw sequencing data in this work has been deposited at the National Genomics Data Center (NGDC; https://ngdc.cncb.ac.cn), China National Center for Bioinformation/ Beijing Institute of Genomics, Chinese Academy of Sciences, with accession number: HRA002496. Data can be accessed via applications, following the GSA guide (https://ngdc.cncb.ac.cn/gsa-human/document). All of the GWAS summary statistics data in this GDBIG platform has been deposited in the Genome Variation Map (GVM) in NGDC, under accession number GVP000003 (https://ngdc.cncb.ac.cn/gvm/getProjectDetail?project=GVP000003). The user can contact the corresponding author to apply for permission to access this data. In addition, the website of Genome Database of Born in Guangzhou Cohort Study (GDBIG) is freely available at http://gdbig.bigcs.com.cn/. Researchers can be accessed through an interactive API, which can be found at http://gdbig.bigcs.com.cn/api.html. Use tutorials are provided on the same webpage to assist users in navigating the platform effectively. The code for extracting information from GDBIG is available on the Github repository(https://github.com/BIGCS-Lab/GDBIGtools) and BioCode project at NGDC (https://ngdc.cncb.ac.cn/biocode/tool/7586). Variant dataset and the GWAS summary statistics for the 14 traits can also be accessed through the GDBIG server (http://gdbig.bigcs.com.cn/), with approval from the Human Genetic Resources Administration of China (HGRAC) (ID: 2022BAT1051). Researchers who are interesting in collaborating with the BIGCS group are encouraged to contact Xiu Qiu and.

## Acknowledgements

This work was supported by Guangzhou Women and Children’s Medical Center (GWCMC). We thank the GWCMC systems team for support of computational and storage resources. We are grateful to all the participants in BIGCS, and the whole BIGCS team. Particularly, we would like to thank Wenzhi Lai, Shunping Liu and Liang Wei from Guangzhou Aixunpan technology Co., Ltd for providing IT technical support to set up the GDBIG website. This work was supported by the National Key R&D Program of China (Grant No. 2024YFC2706801); the National Natural Science Foundation of China (Grant No. 32470679, 81673181, 82173525, and 31900487); the Department of Science and Technology of Guangdong Province Research Foundation (Grant No. 32470679, 2022B1515120080, 2022B1212010004, 2020A1515110859, 2020B1111170001, 2019B020227001, and 2019B030301004); Natural Science Foundation of Guangdong Province (Grant No. 2022B1515120080).

## Supplementary material

Supplementary information to this study can be found online at:

