## Supplementary information for "GDBIG: the first birth cohort genomic database and platform facilitating intergenerational genetic research"

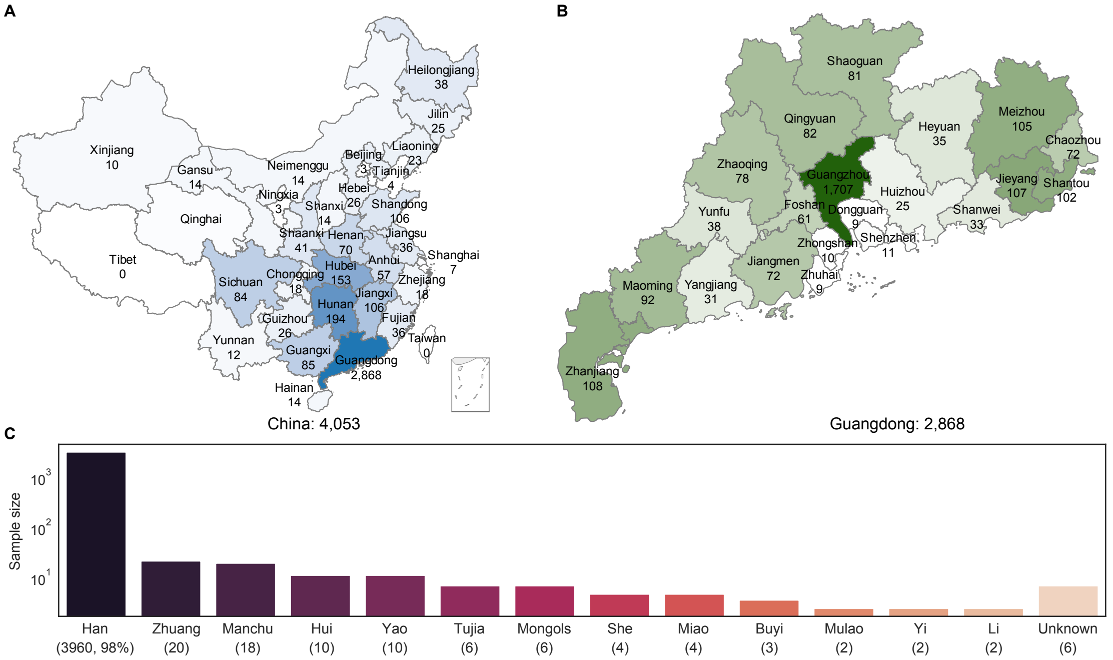


**Figure S1. Geographical and ethnic distribution of the 4,053 BIGCS Phase I samples in GDBIG.** (A) Geographical distribution of the 4,053 participants across China. The participants were assigned to provinces based on their identity records that reflects their birth place. Colors represent the sample size, with darker shades indicating larger sample sizes. (B) Geographical distribution of participants born in Guangdong province. (C) Ethnic distribution of the participants. The y-axis represents the sample size on a log-scale, and each color bar on the x-axis represents an ethnic group, with sample size indicated in parentheses.
